# The Majority of SARS-CoV-2 Plasma Cells are Excluded from the Bone Marrow Long-Lived Compartment 33 Months after mRNA Vaccination

**DOI:** 10.1101/2024.03.02.24303242

**Authors:** Doan C. Nguyen, Ian T. Hentenaar, Andrea Morrison-Porter, David Solano, Natalie S. Haddad, Carlos Castrillon, Pedro A. Lamothe, Joel Andrews, Danielle Roberts, Sagar Lonial, Ignacio Sanz, F. Eun-Hyung Lee

**Affiliations:** Division of Pulmonary, Allergy, Critical Care, and Sleep Medicine, Department of Medicine, Emory University, Atlanta, GA, United States; Division of Rheumatology, Department of Medicine, Emory University, Atlanta, GA, United States; Department of Hematology and Medical Oncology, Winship Cancer Institute, Emory University, Atlanta, GA, United States; Lowance Center for Human Immunology, Emory University, Atlanta, GA, United States

## Abstract

The goal of any vaccine is to induce long-lived plasma cells (LLPC) to provide life-long protection. Natural infection by influenza, measles, or mumps viruses generates bone marrow (BM) LLPC similar to tetanus vaccination which affords safeguards for decades. Although the SARS-CoV-2 mRNA vaccines protect from severe disease, the serologic half-life is short-lived even though SARS-CoV-2-specific plasma cells can be found in the BM. To better understand this paradox, we enrolled 19 healthy adults at 1.5-33 months after SARS-CoV-2 mRNA vaccine and measured influenza-, tetanus-, or SARS-CoV-2-specific antibody secreting cells (ASC) in LLPC (CD19^−^) and non-LLPC (CD19^+^) subsets within the BM. All individuals had IgG ASC specific for influenza, tetanus, and SARS-CoV-2 in at least one BM ASC compartment. However, only influenza- and tetanus-specific ASC were readily detected in the LLPC whereas SARS-CoV-2 specificities were mostly excluded. The ratios of non-LLPC:LLPC for influenza, tetanus, and SARS-CoV-2 were 0.61, 0.44, and 29.07, respectively. Even in five patients with known PCR-proven history of infection and vaccination, SARS-CoV-2-specific ASC were mostly excluded from the LLPC. These specificities were further validated by using multiplex bead binding assays of secreted antibodies in the supernatants of cultured ASC. Similarly, the IgG ratios of non-LLPC:LLPC for influenza, tetanus, and SARS-CoV-2 were 0.66, 0.44, and 23.26, respectively. In all, our studies demonstrate that rapid waning of serum antibodies is accounted for by the inability of mRNA vaccines to induce BM LLPC.

## Introduction

As of December 2023, SARS-CoV-2 (SARS2) has infected over 772 million people worldwide and killed seven million, including 1.2 million in the United States alone^1^. While the original wildtype SARS2 primary vaccine series and boosters have been effective against severe disease, hospitalization, and death, protection with sterilizing immunity against infection or transmission has not been entirely evident. SARS2 vaccines appear to provide lasting T cell responses. However, waning neutralizing antibody levels within 3-6 months results in notable breakthrough infection (BTI) or reinfections with the same strain^2–4^. Therefore, we asked whether subjects after SARS2 vaccination develop SARS2 spike-specificity in the long-lived plasma cell (LLPC) subset (CD19^−^CD38^hi^CD138^+^) of the human bone marrow (BM)^5^. For clarity, the term ASC refers to all antibody-secreting cells (ASC), which include early-minted ones (oftentimes referred to as plasmablasts) and more mature ASC known as plasma cells that can contain LLPC.

Early in the pandemic, reports of SARS2 spike-specific IgG ASC were readily identified in the BM after SARS2 infection or vaccination^6,7^, or in the non-human primates after SARS2 spike protein vaccination^8^, suggesting long-lived humoral protection without serologic confirmation. Interestingly, BM ASC compartments are quite heterogeneous comprising of early-minted ASC (new arrivals) of which some progressively mature into LLPC^9–14^. How LLPC are generated is not entirely clear but after vaccination, the majority of ASC released from secondary lymph nodes are destined to undergo apoptosis unless they finally arrive in the specialized BM survival niches filled with mesenchymal stromal cells and myeloid cells, which provide important factors for survival and maturation such as IL-6 and APRIL^15,16^. These new arrivals can further differentiate into a mature long-lived phenotype, LLPC (CD19^−^CD138^+^), which secrete neutralizing antibodies for decades^9,10^. Although the human BM is a reservoir of LLPC, new arrivals, including CD19^+^CD138^−^ and intermediate phenotypes of CD19^+^CD138^+^ ASC, make it quite heterogenous^17^, such that a mere presence to this locale may not assign durability.

Tetanus vaccination generates antigen (Ag)-specific BM LLPC and affords safeguards for decades with a serologic half-life of 10 years^9,18^. For influenza, humoral immune protection provided by influenza vaccines typically wanes within 4-6 months^19^; however, natural infection provides long-lasting immunity as shown when elderly adults maintained neutralizing antibodies to the 1918 Spanish influenza virus nearly 90 years after the primary infection^20^. Infants may have preexisting maternally derived anti-influenza antibodies although they wane over the first 6 months of life^21^. Unvaccinated individuals are estimated to have their first influenza infection within 5 years of birth^22^ and to be infected with a new influenza virus strain every 3-7 years^23^. Furthermore, newly induced immune responses are enhanced owing to cross-reactive antibodies from infections and reinfections with antigenically similar influenza virus strains^24–28^. Although influenza vaccine induces short-lived protection, natural infection to influenza viruses generates long-lasting humoral immunity to the infecting strain^19,20^.

Here, we measure SARS2 spike-specific ASC in multiple BM compartments up to 33 months after vaccination and compare them to well-known long-lived responses such as tetanus- and influenza-specific ASC by ELISpots. We also validate these results from the cultured BM ASC with secreted antibodies in the supernatants with new bead-based methods. Both assays show that SARS2 IgG responses are excluded from the BM LLPC compartment as early as two months and up to nearly three years after immunization, although they are readily detectable in the non-LLPC subsets of the human BM (CD19^+^CD138^−^ and CD19^+^CD138^+^). In contrast, from the same individuals, tetanus- and influenza-specific ASC are easily detected in both the BM CD19^+^ (non-LLPC) and CD19^−^ (LLPC) compartments. In all, unlike long-lived specificities, SARS2-specific ASC are mainly excluded from the BM LLPC in healthy subjects, despite being present in mature CD19^+^CD138^+^ ASC. This finding provides a mechanistic explanation for the short duration of antibody responses to SARS2 mRNA vaccines. Together with our previous demonstration that LLPC mature from earlier BM ASC precursors, this study provides an experimental model to identify the requirements for full differentiation of BM LLPC.

## Results

### Demographic and clinical characteristics of the 19 BM subjects

From May 2021 until October 2023, we enrolled 19 healthy adults aged of 20-69 years old (**Fig. 1a**). The subjects were recruited for BM aspirates 1.5-33 months after receiving the first dose of SARS2 mRNA vaccines. All received a total of 2 to 5 vaccine doses, and BM aspirates were obtained 0.5-21 months after receiving the last booster (the third, the fourth, or the fifth dose) (**Table 1**). One subject provided 3 longitudinal BM samples over a period of 21 months, resulting in a total of 21 BM aspirates. Five subjects reported infection with SARS2 2-17 months prior to the BM collection, of which two subjects had infection once and three had two subsequent PCR proven SARS2 infections. These infections occurred 1-15.5 months after receiving the most recent vaccine dose (either the first, the second, or the third booster dose). Two of these 5 subjects had infections before vaccination (hybrid immunity) and post-vaccination infection was ruled out by symptomatic cases proven by rapid antigen or PCR testing in all other subjects. All 19 individuals received the quadrivalent influenza vaccine within 1-12 months (relative to the time of each BM aspirate) and one was delayed 1 year due to the pandemic. All received the childhood series of the tetanus toxoid vaccine with recent boosters ranging from one month to 24 years from the time of BM aspirates.

**Fig. 1.**
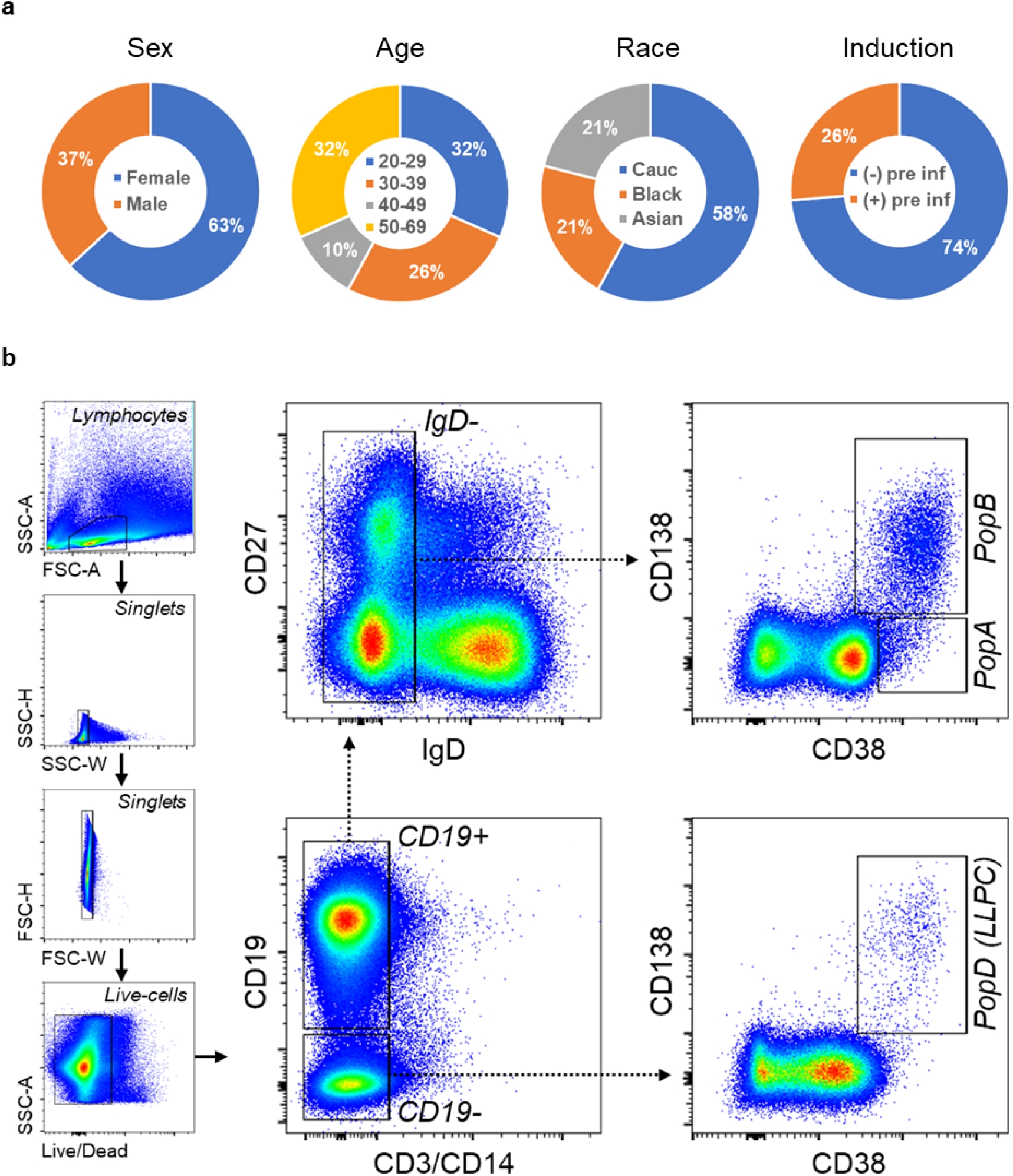
Demographics of the 19-BM-subject cohort. Age, years; Cauc, Caucasian; pre inf, previous (COVID-19) infection (**a**). **General FACS gating strategy used for sorting BM ASC subsets**. BM MNC were first gated for lymphocytes, singlets, and viable cells (based on their FSC/SSC and Live/Death properties) (**b**). CD3 and CD14 were then used as dump markers to capture CD19^+^ and CD19^−^ B cell populations. Subsequent sub-gating from CD19^+^ population on the IgD^−^ fraction (versus CD27) and using CD138 versus CD38 allow for breaking down BM ASC populations into 3 subsets of interest: PopA (CD19^+^CD38^hi^CD138^−^), PopB (CD19^+^CD38^hi^CD138^+^), and PopD (LLPC; CD19^−^CD38^hi^CD138^−^).

**Table 1.**
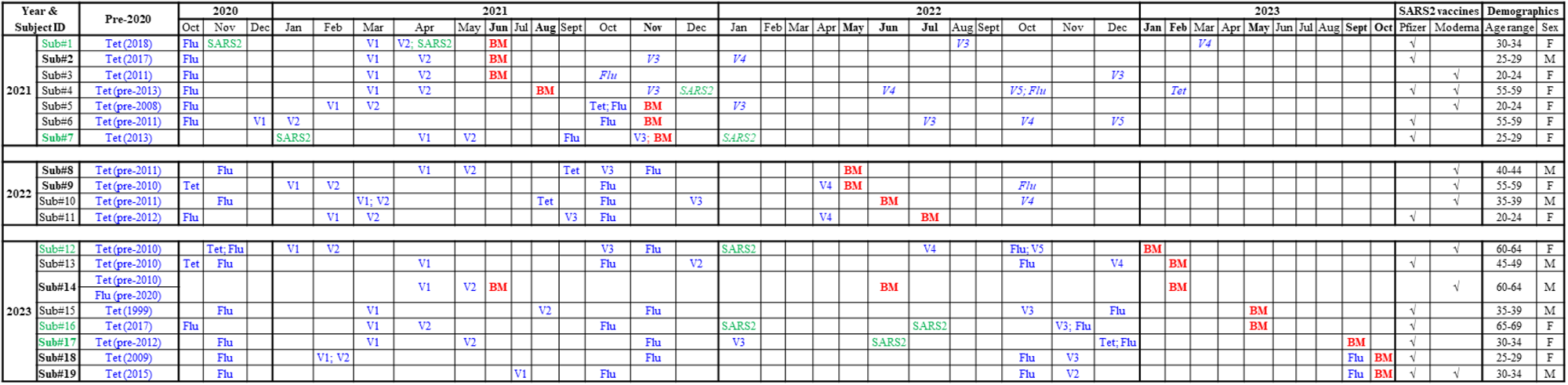
BM subjects and BM samples. During May 2021-October 2023, 19 healthy adults aged of 20-69 years old who were 1.5-33 months after receiving the first dose of SARS2 mRNA vaccines were recruited for BM aspirates. All subjects received the quadrivalent influenza vaccine within 1-12 months (relative to the time of each BM aspirate) and one was delayed 1 year due to the pandemic. All individuals received the childhood series of the tetanus toxoid vaccine with recent boosters ranging from one month to 24 years from the time of BM aspirates. Abbreviation: Sub, subject; Flu, influenza; Tet, tetanus toxoid; SARS2, SARS-CoV-2; V, SARS-CoV-2 mRNA vaccine dose; BM, bone marrow; F, female; M, male. Color and text codes: (blue text) Vaccination (Tet, Flu, or SARS2); V1-V5, dose 1-5 of SARS2 vaccination; (green text) Infection (with SARS2); (**bold text**) **Sub#**, with multiplex bead binding assays performed (on the cultured supernatants); (**bold text, month**) **Jun**, month with BM aspirates collected; (*italic text*), occurred after BM aspiration. For the multiple-aspirate subject (i.e. Sub#14), age indicated at the most recent BM sample collection.

### BM non-LLPC (PopA and PopB) and LLPC (PopD) subsets and antigen validation

BM ASC subsets were FACS-sorted according to surface expression of CD19, CD38, and CD138, as previously described^9^ (**Fig. 1b**). To overcome the problem with the rapid death of ASC *ex vivo*^15,16^, we rested the ASC overnight in a new human *in vitro* plasma cell survival system which is capable of maintaining ASC viability for months^15^. Since we had previously localized the BM LLPC compartment into PopD (CD19^−^CD38^hi^CD138^+^)^9,16^, this population was sorted out of total BM ASC together with PopA (CD19^+^CD38^hi^CD138^−^) and PopB (CD19^+^CD38^hi^CD138^+^), and tested for total IgG secretion as well as SARS2-, influenza-(Flu-), and tetanus toxoid-(Tet-) specific IgG secretion by bulk ELISpots. To optimize antigen detection for the BM assays, we collected early-minted blood ASC (CD27^hi^CD38^hi^; **Suppl. Fig. S1**) 6-7 days after SARS2, Tet, or Flu vaccine, which is the peak time for enrichment of vaccine-specific ASC in the blood after secondary immunization^29,30^, and performed ELISpots (**Suppl. Fig. S2a**). Of the SARS2 antigens (S1, S2, RBD, S2P, NTD, and NP proteins), S2P, a prefusion-stabilized SARS2 spike trimer^31^, generated the highest frequency, followed by S1 (with no significant difference; *p*=0.21) (**Fig. 2a,b**), and so S2P was selected for BM ELISpot assays. We also validated the quadrivalent Flu vaccine (seasons of 2019-20 to 2023-24) and Tet antigen (**Suppl. Fig. S2b,c**), which are enriched in blood ASC at the peak of the respective vaccines^29,30^.

**Fig. 2.**
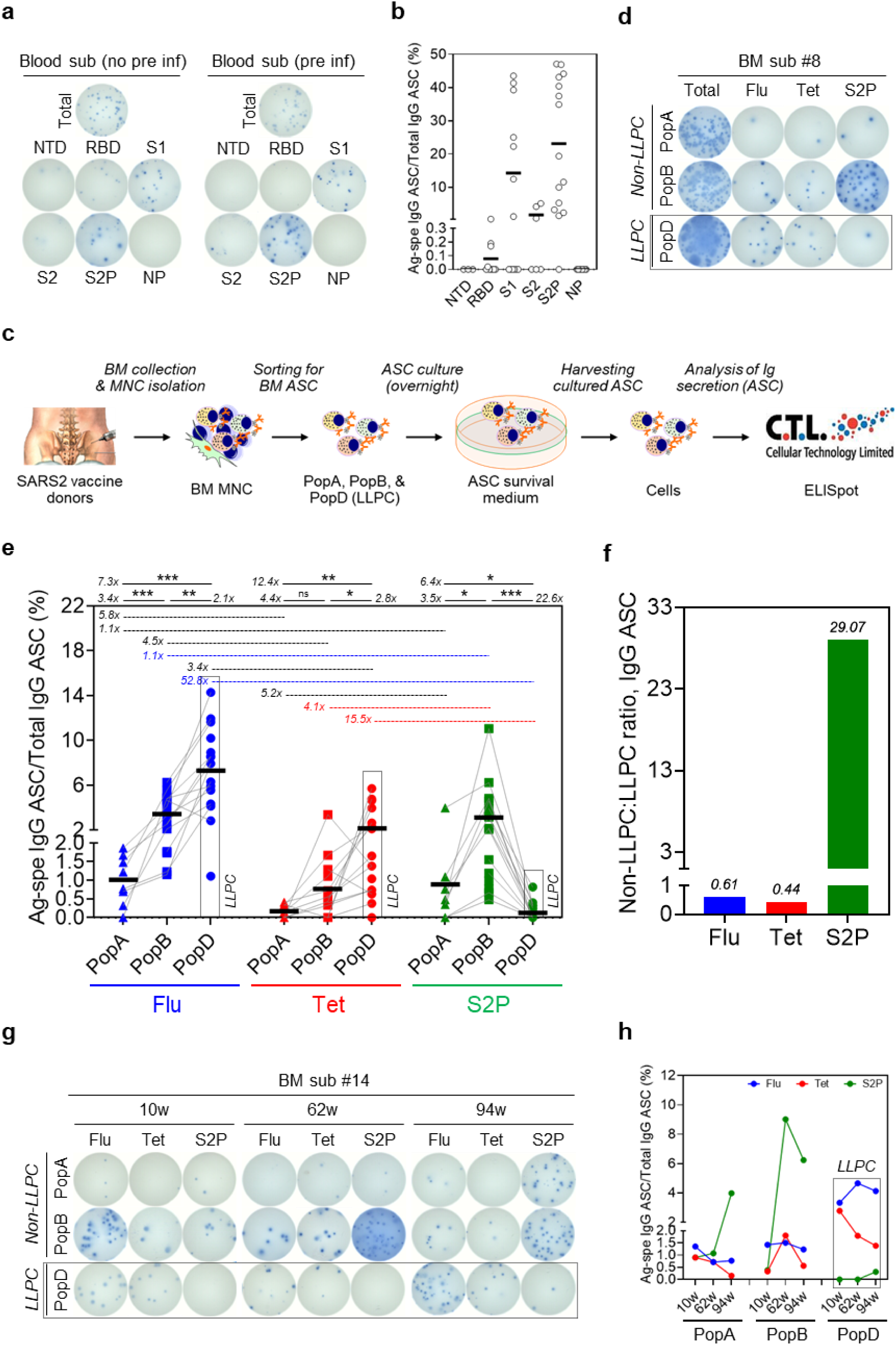
Exclusion of SARS-CoV-2-specific BM IgG LLPC after SARS2 mRNA vaccines by ELISpots. The S2P antigen is most sensitive to capture SARS-CoV-2-specific blood ASC isolated after SARS2 mRNA vaccines. Blood ASC from subjects at the peak time which is 5-7 days post-vaccine (**a**,**b**). Representative ELISpot scanned images (left, a vaccinated subject without previous COVID-19 infection; right, a vaccinated subject with previous COVID-19 infection) (**a**). The numbers of input ASC that were incubated for total IgG ∼687 (left) or ∼522 (right), and antigen-specific ASC IgG ∼2,062 (left) or ∼1,566 (right). Frequencies (%) of Ag-specific IgG ASC per total IgG ASC. Each circle represents an individual vaccine subject (**b**). For additional Ag validation, see **Suppl. Fig. S2**. Summary of the techniques and the experimental design for total, Flu, Tet, and S2P-specific ASC by ELISpots (**c**). Total IgG, and antigen-specific ASC from PopA, PopB, and PopD for Flu-, Tet-, S2P-specific BM ASC. Representative ELISpot scanned images shown (**d**). The numbers of input ASC that were incubated were ∼52K, ∼12.1K, and ∼10.1K for PopA, PopB, and PopD, respectively. Each symbol represents an individual vaccine subject for total IgG and antigen-specific ASC from PopA, PopB, and PopD for Flu-, Tet-, S2P-specific BM ASC (**e**). The numbers indicate fold difference (i.e. ratios) when comparing different BM ASC subsets for the same Ag specificity or comparing different Ag specificities within the same BM ASC subset. ns, not significant; *p<0.05; **p<0.005; ***p<0.0005. Fold difference (i.e. ratios) when comparing different Ag specificities between non-LLPC (i.e. combined PopA and PopB) versus LLPC (PopD) (**f**). BM IgG ASC response kinetics in the SARS2 mRNA vaccinee who donated three longitudinal BM aspirates (**g**,**h**). Representative ELISpot scanned images. The numbers of input ASC that were incubated were ∼21K, ∼40K, and ∼4.9K (*10w*); ∼14K, ∼12K, and ∼3.8K (*62w*); and ∼58K, ∼22K, and ∼7.2K (*94w*) for PopA, PopB, and PopD, respectively (**g**). Ag-specific BM IgG ASC response kinetics (**h**). Sub, subject; K: 1,000; LLPC: long-lived plasma cell (dotted boxes in **d**, **e**, **g**, and **h**); Flu: influenza; Tet: tetanus; Pre inf: previous (COVID-19) infection; d: day; w: week; m: month; y: year. For details of subjects and samples, see **Table 1**.

### Exclusion of SARS2-specific BM IgG PopD as measured by ELISpots

Since BM aspirates can yield variable cell numbers, we included BM aspirates with >3,000 sorted cells in each of the three ASC populations and performed bulk ELISpots (**Fig. 2c**). Among the 21 BM samples, sufficient cells to confidently measure vaccine-specificities within PopA, PopB, and PopD were obtained from 8, 15, and 17 samples. As previously shown, all BM ASC subsets had detectable total IgG ASC. Similar to previous reports^9^, PopD contained the highest percentage of Flu- and Tet-specific IgG ASC per total IgG ASC: mean 7.3% (7.31±3.51) and 2.1% (2.14±1.70) respectively (**Fig. 2d,e**). PopB were readily populated with Flu- and Tet-specific IgG ASC: mean 3.4% (3.43±1.68) and 0.8% (0.77±0.87) respectively, and with the lowest in PopA: mean 1% (1.0±0.66) and 0.2% (0.17±0.17) respectively. Strikingly, within the same subjects, we could rarely detect S2P-specific ASC in PopD: mean 0.1% (0.14±0.23). In contrast, the S2P specificity was readily found in PopB and PopA at frequencies comparable to Tet and Flu: mean 3.1% (3.13±2.82) and 0.9% (0.89±1.3) respectively. Although frequencies of Tet-specific ASC in PopA versus PopB showed no statistically significant difference, frequencies of Flu-specific IgG were higher in PopB over PopA. For both Flu- and Tet-specific ASC, the frequencies in PopD were always higher than in PopB. In contrast, the SARS2 ASC frequencies were always significantly lower in PopD compared to PopB (**Fig. 2d,e**). On average, the fold changes of IgG ASC specificities within PopD were 52.8 times for Flu:SARS2 and 15.5 times for Tet:SARS2 (**Fig. 2e**). In comparison, the fold changes of IgG ASC specificities within PopB were 1.1 fold for Flu:SARS2 and 4.1 fold for Tet:SARS2. Overall, the ratios of non-LLPC:LLPC for Flu-, Tet-, and SARS2 were 0.61, 0.44, and 29.07, respectively (**Fig. 2f**).

### Kinetic responses for IgG ASC in a longitudinal BM aspirate collection

We next assessed the IgG ASC kinetic response in a single subject who provided 3 sequential BM aspirates over a period of 23 months after the first vaccine. BM aspirates were taken 10, 62, and 94 weeks after the first SARS2 vaccine dose (or 6, 58, and 90 weeks after the second vaccine dose) (**Table 1**). Each BM aspirate provided >3,000 FACS sorted ASC in each subset. Again, total IgG ASC were detected in all BM PopA, PopB, and PopD. We observed an increase in the frequencies of S2P-specific IgG ASC in PopA and PopB at 62 weeks (1.07% and 9.02% respectively) and 94 weeks (3.98% and 6.24% respectively) compared to the first time point (0.90% and 0.38%, respectively) (**Fig. 2g,h**). However, in PopD, there were no S2P-specific IgG spots detected at the first two time points and only 0.31% at 94 weeks post-vaccination. Notably, at the earliest time-point (10 weeks), the highest S2P-specific IgG ASC frequency was observed in PopA which then increased in PopB at 62 and 94 weeks. In all, regardless of time points, the S2P-specific ASC frequencies were always higher in PopA and PopB compared to PopD, which was quite rare even at 94 weeks. As expected, we observed the highest Flu- and Tet-specific frequencies in PopD, followed by PopB, and lowest in PopA (**Fig. 2g,h**). Interestingly, the Flu- and Tet-specific frequencies were quite consistent over the course of two years in these BM subsets.

### Exclusion of SARS2-specific BM IgA PopD

Similar to IgG ASC, the frequency of Flu- and Tet-specific IgA ASC was highest in PopD with a mean of 1.7% (1.70±0.45) and 0.3% (0.31±0.12) respectively, while PopA and PopB were lower: for Flu, mean 0.8% (0.82±0.43) and 1.4% (1.35±1.32) respectively, and for Tet, 0.2% (0.24±0.34) and 0.1% (0.11±0.10) respectively (**Suppl. Fig. S3a,b**). Consistent with previous studies^32^, these results may be explained by the predominance of IgG responses to the intramuscular tetanus vaccine. Similar to IgG ASC, S2P-specific IgA ASC were also detected predominantly in PopA and PopB with a mean of 1.5% (1.46%±1.57) and 0.9% (0.90±0.66) respectively, and was virtually absent in PopD: a mean of 0.03% (0.03%±0.06) (**Suppl. Fig. S3**). Thus, similar to IgG ASC, other class switched isotypes such as S2P-specific IgA ASC are also excluded from PopD.

### Exclusion of SARS2-specific IgG secreted in PopD as measured in culture supernatant

Next, to validate the antigen-specific ELISpot responses, we measured secreted IgG from BM ASC subsets using a novel plasma cell culture system (**Fig. 3a**; see also Methods). Briefly, from the 8 individuals that yielded sufficient sorted cells, we cultured BM ASC subsets (PopA, PopB, and PopD) in a specialized *in vitro* BM mimetic system overnight^15^ and measured the cultured supernatants for IgG specific for Flu, Tet, and S2P by multiplex bead-binding assays (Luminex)^33^. The results were similar to the ELISpot numbers (**Fig. 3b**). The percentages of Flu- and Tet-IgG per total IgG were highest in PopD (mean 7.92±7.41 and 7.51±9.98 respectively) compared to PopB (mean 4.09±2.81 and 2.30±2.14 respectively) or PopA (mean 1.12±1.08 and 0.97±2.46 respectively). In contrast, the percentage of S2P-IgG per total IgG was lower in PopD (mean 0.12±0.20) compared to PopA (mean 0.31±0.62) and especially PopB (mean 2.46±1.83). Of the 8 individuals, the fold change in PopD for Flu:SARS2 was 66.5 and for Tet:SARS2 was 63.1 (**Fig. 3b**). In comparison, the fold change within PopB for Flu:SARS2 was 1.7 and for Tet:SARS2 was 0.9, demonstrating similar quantities of IgG to Flu, Tet, and SARS2 in PopB. Ultimately, using this method of measuring secreted antibodies from the cultured BM ASC, the ratios of non-LLPC:LLPC for Flu-, Tet-, and SARS2 from BM ASC culture supernatant were 0.66, 0.44, and 23.26, which was similar to the ELISpot results (**Fig. 3c**). In all, we validated the antigen specificities observed by the ELISpots using our novel *in vitro* plasma cell culture method which also showed exclusion of SARS2-specific ASC in PopD.

**Fig. 3.**
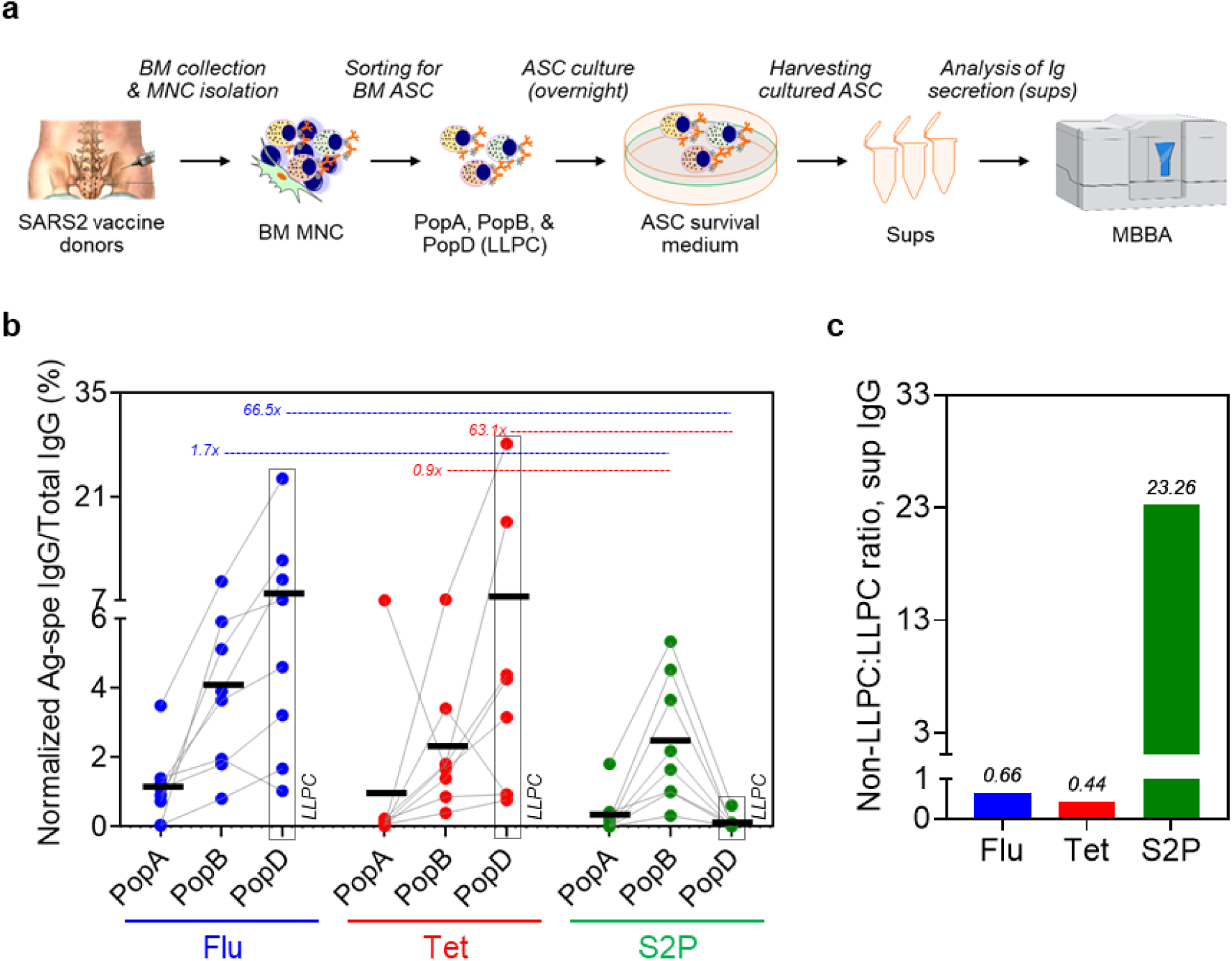
Exclusion of SARS-CoV-2-specific BM IgG LLPC in SARS2 mRNA vaccinees by IgG detection from the BM ASC culture supernatants. Summary of the techniques and the experimental design (**a**). From the cultures of BM ASC, the supernatant preps were collected for quantitation of Ag-specific IgG. Sups, BM ASC culture supernatant preps; MBBA, multiplex bead binding assay. MBBA measuring IgG specific for Flu, Tet, and S2P (normalized to total IgG and ASC input numbers) from culture supernatant of PopA, PopB, and PopD of 8 individuals (**b**). All supernatants were collected for 18-24 hours in culture from BM ASC during revival from the FACS sorters and were tested undiluted (neat). Fold difference (i.e. ratios) when comparing normalized Ag-specific IgG in the supernatants from the culture of non-LLPC (combined PopA and PopB) versus LLPC (**c**). For ratio calculation, see **Methods**. For IgG standard vs MFI curve, see **Suppl. Fig. S4**. LLPC: long-lived plasma cell (dotted boxes in **b**); Flu: influenza; Tet: tetanus. For details of subjects and samples, see **Table 1**.

### Only 35% of subjects with SARS2-specific ASC in PopD

Finally, we calculated the number of individuals with S2P-positive responses for each BM ASC subset. S2P IgG ASC were easily detected in PopA in 6/8 (75%) individuals and in PopB, in all 15/15 (100%) subjects (**Fig. 4a**). Only 6/17 (35.29%) subjects had S2P IgG ASC in PopD, and all were extremely low frequencies despite four or five doses of the vaccine and multiple known SARS2 infections. As expected, nearly all subjects had easily detectable Flu- and Tet-specificities in PopD: 17/17 (100%) and 16/17 (94.12%) respectively. Altogether, durable serologic immune response correlates well with the abundance of Flu- and Tet-specific ASC in PopD while short-lived serologic antibody responses to SARS2 mRNA vaccines may explained by exclusion of SARS2-specific ASC from this BM compartment (summarized in **Fig. 4b**).

**Fig. 4.**
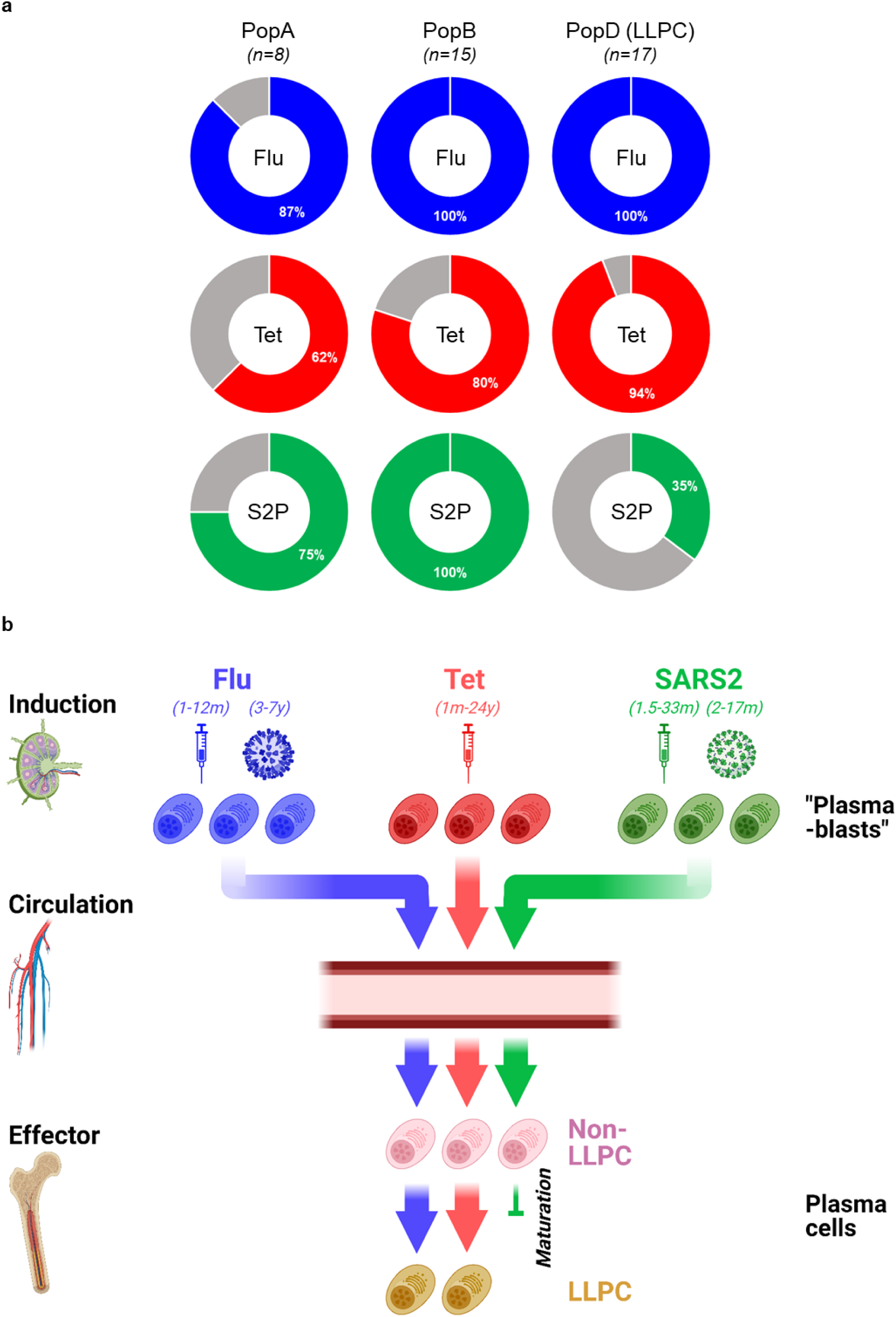
Antigen specificity strata of all individuals examined for each BM ASC subset. *n*, number of BM donors (**a**). Graphical summary. The majority of SARS2 plasma cells are excluded from the BM LLPC compartment 33 months after mRNA vaccination (**b**). Created with BioRender.com.

## Discussion

In this study, we show that although there is an abundance of SARS2-specific ASC in the BM short-lived plasma cell compartment, these cells are largely excluded from the LLPC compartment. This phenomenon is in stark contrast to influenza- and tetanus-specificities which are inherent to the BM LLPC. Hence, the lack of SARS2-specific ASC in the LLPC provides a mechanistic explanation of the short-lived serology of mRNA vaccines and how these mRNA platforms are unable to induce the LLPC formation. Early-minted ASC typically journey to the BM microniches and require time to fully mature into LLPC. Fundamental changes of the new ASC arrivals are required for maturation into LLPC. These BM maturation programs where an early-minted ASC undergoes dramatic morphological, transcriptional, and epigenetic modifications together with metabolic alterations provide the final maturation steps to become a LLPC^10^. Increased Ig transcripts^11^, increased unfolded protein response (UPR)^34^, and anti-apoptotic^10^ and autophagy^35^ programs are just a few of the pathways involved in ASC maturation^36^. Because this progression is arduous, not all the new arrivals can ultimately complete this entire LLPC process. Moreover, the exact kinetics of this maturation also remain unknown. In all, our study offers major insights into how the mRNA vaccines fail to induce the necessary ASC precursors with programs that are required to fully mature into LLPC.

The exact mechanisms of how LLPC are formed are still under investigation. At one time, it was thought that all human ASC had the potential to become LLPC by simply migrating to environments rich in survival factors; but recent evidence shows how imprinting of an early-minted ASC at the time of priming in addition to terminal maturation in survival niches endow particular properties for durability. LLPC are thought to come from memory B cells (mBC)^37^. Thus, a longer interval between prime-boost vaccine strategies for mBC formation may play a role in the generation of LLPC. Also, the phenotype of mBC such as FcRL5^+^ and the cytokine milieu i.e. IFNγ may also play key roles in the generation of LLPC^38^. It is likely that imprinting at the time of B cell induction also determines the LLPC fate but with SARS2 mRNA vaccines, they fail to imprint these LLPC programs. Finally, even after 33 months after the vaccine, we show that SARS2-specfic ASC are still excluded from the BM LLPC compartment. Thus, a longer tincture of time is unlikely to fill the LLPC subset.

In the patient with sequential BM aspirates nearly two years after the initial vaccine, there are two explanations for the abundant S2P specificity in PopA at 94 weeks and PopB at 62 and 94 weeks. Conventionally, PopA and PopB are considered the result of more recent immune responses. Thus, the very high frequency in PopA and PopB suggests another exposure to SARS2 antigens such as breakthrough asymptomatic infections temporally close to the corresponding BM sampling. Asymptomatic infections have been well described with the emergence of the highly transmissible Omicron variants^23^. Thus, assuming an asymptomatic infection, even 94 weeks after vaccination and infection(s), S2P-specific ASC cannot fill the LLPC compartment. Alternatively, it could be argued that lymph node S2P-specific IgG ASC take time to migrate to the BM microenvironment and reflect the product of ongoing germinal center (GC) reactions, which have been documented to last for up to 6 months after vaccination which is well after the peak of early-minted ASC responses in the blood (day 7)^7^. This argument still emphasizes the fact that even two years after the vaccines, PopB cannot differentiate into LLPC even with ongoing persistent GC.

Our results are consistent with recent BM studies by Tehrani et al. demonstrating that most spike-specific ASC are detectable in the CD19^+^ compartments post-SARS2 infection alone^39^. However, in this study, BM sampling was collected only 5-8 months post-illness and not up to 3 years as in our study^39^. Most importantly, this study used frozen BM ASC which are less reliable due to the fragility of BM ASC populations upon thawing. Also, the authors did not include longitudinal samples, IgA isotypes, Flu specificity, or PopA. Nonetheless, 5-8 months after infection alone, SARS2-specific ASC were also excluded in LLPC, similar to the finding in our study after vaccination.

In another flow cytometry-based BM study, Schulz et al. also found S1-specific responses predominately in the BM CD19^+^ compartment after vaccination^40^. In this study, BM samples were collected during hip joint replacement surgery from patients of up to 17 months post-SARS2-vaccination. While no Flu- or Tet-specificities were evaluated, the authors noted some SARS2 specificity in the CD19^neg^ ASC compartment and concluded they were in fact long-lived, but these SARS2 specific ASC were notably in the CD45^+^ (of CD19^neg^) ASC subset^40^. The majority of LLPC as originally defined in Halliley et al. demonstrate downregulated CD45 in our LLPC^9^ (**Suppl. Fig. S5**), which is also consistent with previous studies^41,42^. Interestingly, in concordance with our study, in Schulz et al, the CD19^neg^CD45^neg^ subset which includes the majority of our previously defined LLPC also excluded the S1-specific responses^40^. Hence, the *bona fide* LLPC which may be a subset of the CD19^neg^ BM ASC population likely harbors Flu- and Tet-specificities as well as measles- and mumps-specificities but appears to exclude SARS2-specific responses.

We cannot rule out a subset of BM PopB cells which may be an intermediary population on the road to maturing into the LLPC. Our previous single-cell transcriptional data showed that the most mature BM clusters with aggregated LLPC also contained some PopB^11^. Thus, simple surface markers CD19 and CD138 may be too blunt an instrument to dissect the heterogeneity of BM PopB. PopB likely contains some new arrivals as well as some early mature BM ASC subsets since Tet-specific responses reside in PopB and LLPC years after vaccination albeit at much lower frequencies in PopB compared to LLPC. Ultimately, additional dissection into the transcriptional and epigenetic differences in Tet-versus S2P-specific PopB (CD19^+^CD138^+^) cells may reveal important mechanistic differences in forming long-lived ASC.

Although the emergence of new viral variants confounded serum protection, in this study, we focused on responses against the original virus and the wildtype vaccines, knowing that they rapidly wane within 3-6 months regardless of the vaccine platform (mRNA and viral vector-based vaccines like adenovirus (Ad) vectors)^3,4^. The unstable nature of mRNA and the resultant transient expression of the spike protein during induction might explain the lack of sustained antibody responses. Interestingly, the Ad vectors persist for weeks, yet specific humoral immunity is also short-lasting^3,4^. Furthermore, recent evidence showed that the mRNA vaccine platforms do not induce strong type I IFN responses^43^. *In vitro* and animal studies suggest that type I IFN may play a role in LLPC^44,45^, but whether it is important for the generation of *in vivo* human LLPC will need further studies. Given both the mRNA and Ad vector vaccine platforms induce strong germinal center reactions and interactions with Tfh cells, the mechanisms underlying their failure to generate LLPC are even more puzzling^2^ suggesting dysfunction in the maturation process in BM microniche. In all, we merely suggest that before moving the current vaccines to the mRNA platforms, more studies on the durability of humoral immunity are needed.

Could the limited durability of neutralizing antibody responses be due to the structural nature of the spike protein itself and thus limited only to coronavirus vaccines? Coronaviruses lack highly repetitive organized structures or pathogen-associated structural patterns^46^. Most RNA viruses that induce long-lasting antibody immunity have on their surface rigid repetitive structures spaced 5-10nm^47^. In coronaviruses, the long spike proteins are embedded in a fluid membrane which are often loosely floating and widely spaced at 25nm apart^46^. Therefore, the inherent nature of the spike protein on coronaviruses itself may be an issue in B cell activation^47^ since neutralizing antibody responses to seasonal human coronaviruses, as well as SARS-CoV-1 and MERS-CoV, are also short-lived^2^. Perhaps the long widely spaced spike protein structure of the coronaviruses may play an important role in antibody durability.

There are limitations in our study. First, our sample size is relatively small especially of those after vaccine and infection. Second, the infections were self-reported symptoms that warranted testing, so any asymptomatic infections were not confirmed. Third, primary BM ASC are rare cell types and BM aspirates are difficult to obtain and interrogate; thus, not all samples provided sufficient cells in each BM subset. Lastly, we had limited longitudinal and sequential samples with the longest at 33 months since the first SARS2 vaccine dose. Of course, it would be important to assess the BM compartment decades after the primary vaccine and as new variant SARS2 viruses continue to circulate.

In conclusion, the holy grail of vaccinology is the generation of LLPC. Our findings demonstrate the exclusion of SARS2 specificity in the BM LLPC compartment and provide novel insights of how the mRNA vaccines fail to induce necessary precursor programs to fully mature into BM LLPC. These findings have implications for the need to improve COVID-19 vaccination. Whether optimizing vaccine regimens or immunization schedules, engineering different spike proteins, or formulating vaccine adjuvants and delivery systems will need better understanding. Finally, interrogating the LLPC require invasive BM aspirates and a greater tincture of time (likely years); thus, identifying early biomarkers of early-minted blood ASC or LLPC precursors are needed to help predict the durability of new platform vaccines.

## Methods

### Healthy human subjects

A total of 21 BM aspirate samples were obtained from 19 healthy adult donors who received Tdap, influenza, COVID-19 primary and booster vaccines. Select demographic and clinical characteristics of the subjects can be seen in **Fig. 1a**. Detail information on BM subjects and BM samples can be found in **Table 1**. For antigen validation, peripheral blood samples were obtained from 64 healthy subjects who received vaccines for Tdap, influenza, or COVID-19 (the 3^rd^, 4^th^, or 5^th^ dose) at 5-7d prior to sample collection. For CD45 flow cytometric staining, an additional of five healthy BM subjects were obtained, acquired, and analyzed. All studies were approved by the Emory University Institutional Review Board Committee.

### Purification of blood and BM ASC

Isolation of peripheral blood mononuclear cells (PBMC) and BM mononuclear cells (BMMC) was performed as previously described^15^. Fresh blood ASC, BM PopA, BM PopB, and BM LLPC (PopD) were purified using FACS-based sorting, as previously described^9,15^. Briefly, mononuclear cells were isolated by Ficoll density gradient centrifugation and enriched by either a commercial human Pan-B cell enrichment kit (that removes cells expressing CD2, CD3, CD14, CD16, CD36, CD42b, CD56, CD66b, CD123, and glycophorin A) (StemCell Technologies) or a custom-designed negative selection cell isolation kit (that removes cells expressing CD3, CD14, CD66b, and glycophorin A) (StemCell Technologies) to limit sorting time and pressure on fragile ASC. Enriched fractions were then stained with the following anti-human antibodies: IgD–FITC (Cat. #555778; BD Biosciences) or IgD-Brilliant Violet 480 (Cat. #566138; BD Biosciences), CD3-BV711 (Cat. #317328; BioLegend) or CD3-BUV737 (Cat. #612750; BD Biosciences), CD14-BV711 (Cat. #301838; BioLegend) or CD14-BUV737 (Cat. #612763; BD Biosciences), CD19-PE-Cy7 (Cat. #560911; BD Biosciences) or CD19-Spark NIR 685 (Cat. #302270; BioLegend), CD38-V450 (Cat. #561378; BD Bioscience) or CD38-Brilliant Violet 785 (Cat. #303530; BioLegend), CD138-APC (Cat. #130-117-395; Miltenyi Biotech) or CD138-APC-R700 (Cat. #566050; BD Biosciences), CD27-APC-e780 (Cat. #5016160; eBiosciences) or CD27-Brilliant Violet 711 (Cat. #356430; BioLegend), and LiveDead (Cat. #L34966; Invitrogen) or Zombie NIR Fixable Viability Kit (Cat. #423106; BioLegend). ASC subsets were FACS-sorted on a BD FACSAria II using a standardized sorting procedure with rainbow calibration particles to ensure consistency of sorts between individuals. ASC subsets were sorted as^9,10,15^: Blood ASC (IgD^−^CD27^hi^CD38^hi^), BM ASC (IgD^−^CD19^+^CD38^hi^), PopA (CD19^+^IgD^−^D38^hi^CD138^−^), PopB (CD19^+^IgD^−^ CD38^hi^CD138^+^), and PopD (CD19^−^IgD^−^CD38^hi^CD138^+^). Sorted ASC populations were generally 93-99% pure (except for PopA whose purity was usually ∼60-75%).

### Antigens for specific Ig immunoassays (ELISpots and multiplex bead binding assays)

The following antigens were used for antigen-specific IgG and IgA capturing: quadrivalent influenza vaccine 2019-20, 2020-21, 2021-22, or 2023-24 (ie for antigen-specific IgG capturing) (Fluarix Quadrivalent Influenza Vaccine 2019-20, 2020-21, 2021-22, or 2023-24 Formula, respectively; GSK Biologicals/ABO Pharmaceuticals; Afluria® Quadrivalent (Seqirus) and Fluzone® Quadrivalent (Sanofi Pasteur)), Tetanus Toxoid, *Clostridium tetani* (Calbiochem/Millipore Sigma or Fina Biosolutions), and SARS-CoV-2 S2P (recombinant SARS-CoV-2 soluble spike trimer protein, lot #P210721.02; Protein Expression Laboratory, Frederick National Laboratory for Cancer Research (FNLCR), Frederick, MD). For relative quantitation of Ag-specific antibody titers, standard curves were generated using monoclonal antibody (mAb) standards of anti-tetanus toxin mAb (clone TetE3; The Native Antigen Company) and SARS-CoV-2-reactive (ie spike RBD) mAb (Abeomics). For determining the concentrations of total IgG, purified human IgG (ChromePure human IgG, JacksonImmuno Research Laboratories) was used as a standard.

### Blood and BM ASC bulk cultures and ELISpot assays

Human ASC cultures were conducted in MSC secretome (ASC survival medium) and in hypoxic conditions (2.5% O2) at 37°C, as previously described^15^. This culture system is called plasma cell survival system (PCSS)^48^. IgG and IgA secretion of cultured ASC was assessed by ELISpot assays, which quantitated IgG- and IgA-secreting cells. These assays used goat anti-human IgG or IgA for total IgG or IgA capturing, respectively, and alkaline phosphatase-conjugated goat anti-human IgG or IgA, respectively, for detection, and were performed as previously described^15^.

### Multiplex bead binding assays (MBBA)

Multiplex bead binding assays (MBBA) were performed on the supernatants collected from culture of BM ASC purified from 8 individuals who provided sufficient post-sort cells for all 3 subsets. For total IgG, biotinylated goat anti-human IgG (Southern Biotech) was conjugated to avidin-coupled MagPlex-avidin microspheres of spectrally distinct regions as previously described^49^. For antigen-specific MBBA, antigens were conjugated to MagPlex microspheres of spectrally distinct regions (Luminex) via standard carbodiimide coupling procedures as previously described^33^. Antigen specific and total IgG MBBA were performed using a Luminex FLEXMAP 3D instrument (Luminex) as previously described^33^. All viral protein coupled microspheres were tested together as a combined multiplex antigen-specific immunoassay; all anti-human immunoglobulin coupled microspheres were tested together as a combined multiplex total Ig immunoassay. Median fluorescent intensity (MFI) using combined or individual detection antibodies was measured using the Luminex xPONENT 4.3 software at enhanced PMT setting. The net MFI was obtained by subtracting the background value. The culture supernatant MFI values were normalized to the relative IgG concentrations (pg/mL) based on the total human IgG standard curves, followed by normalization of these resultant IgG concentrations (pg/mL) to the ASC input numbers and duration of culture (day). The MFI normalization was performed based on the equation as shown in **Suppl. Fig. S4**. Data were expressed as the percents or ratios of the titers of Ag-specific IgG to those of total IgG. All the supernatants were collected after 1 day in culture of off-sorter BM ASC subsets and were tested undiluted (neat) or 1:2 diluted – except for the total IgG titrations which were also assayed at further dilutions (e.g. 1:4, 1:8, 1:16, 1:32, 1:64, 1:128, and 1:256).

### CD45 BM ASC flow cytometry

BMMC were stained with the following anti-human antibodies: IgD-Brilliant Violet 480 (Cat. #566138; BD Biosciences), CD3-BUV737 (Cat. #612750; BD Biosciences), CD14-BUV737 (Cat. #612763; BD Biosciences), CD19-Spark NIR 685 (Cat. #302270; BioLegend), CD38-Brilliant Violet 785 (Cat. #303530; BioLegend), CD138-APC-R700 (Cat. #566050; BD Biosciences), CD27-Brilliant Violet 711 (Cat. #356430; BioLegend), CD134 (OX40)-Brilliant Violet 510 (Cat. #350025; BioLegend), CD246 (ALK)-Alexa Fluor 488 (Cat. #NBP3-08771AF488; Novus), CD357 (GITR)-Brilliant Violet 605 (Cat. #747664; BD Biosciences), and CD45-PE-Cy5 (Cat. #304009; BioLegend). Samples were run on a Cytek’s Aurora Spectral Flow Cytometer analyzed with the FlowJo v10.8.1 software (FlowJo, LLC).

### Statistics

All statistics were assessed using Student’s t-test (two-tailed unpaired t-test) or one-way ANOVA performed with GraphPad Prism (v8.4.2; GraphPad Software). Data are presented as mean±SD and differences were considered significant at *p* values less than 0.05.

## Data Availability

All data produced in the present study are available upon reasonable request to the authors.

## Author Contributions

Conceptualization: DCN and FEL. Methodology: DCN, ITH, AMP, DS, NSH, CC. Resources and sample acquisition: PAL, JA, DR, SL, IS, and FEL. Funding acquisition: FEL. Supervision: FEL. Writing – original draft: DCN and FEL. All authors have reviewed, edited, and approved the final manuscript.

## Acknowledgements

We thank Shuya Kyu, Monica Cabrera-Mora, and Martin C. Runnstrom for technical assistance. We thank Robert E. Karaffa, Kametha T. Fife, and Sommer Durham of the Emory University School of Medicine Flow Cytometry Core for technical support. We thank Rahul Patel, Mindy Hernandez, and our clinical coordinators and the donors who made this study possible.

## Supplemental figures

**Suppl. Fig. S1.**
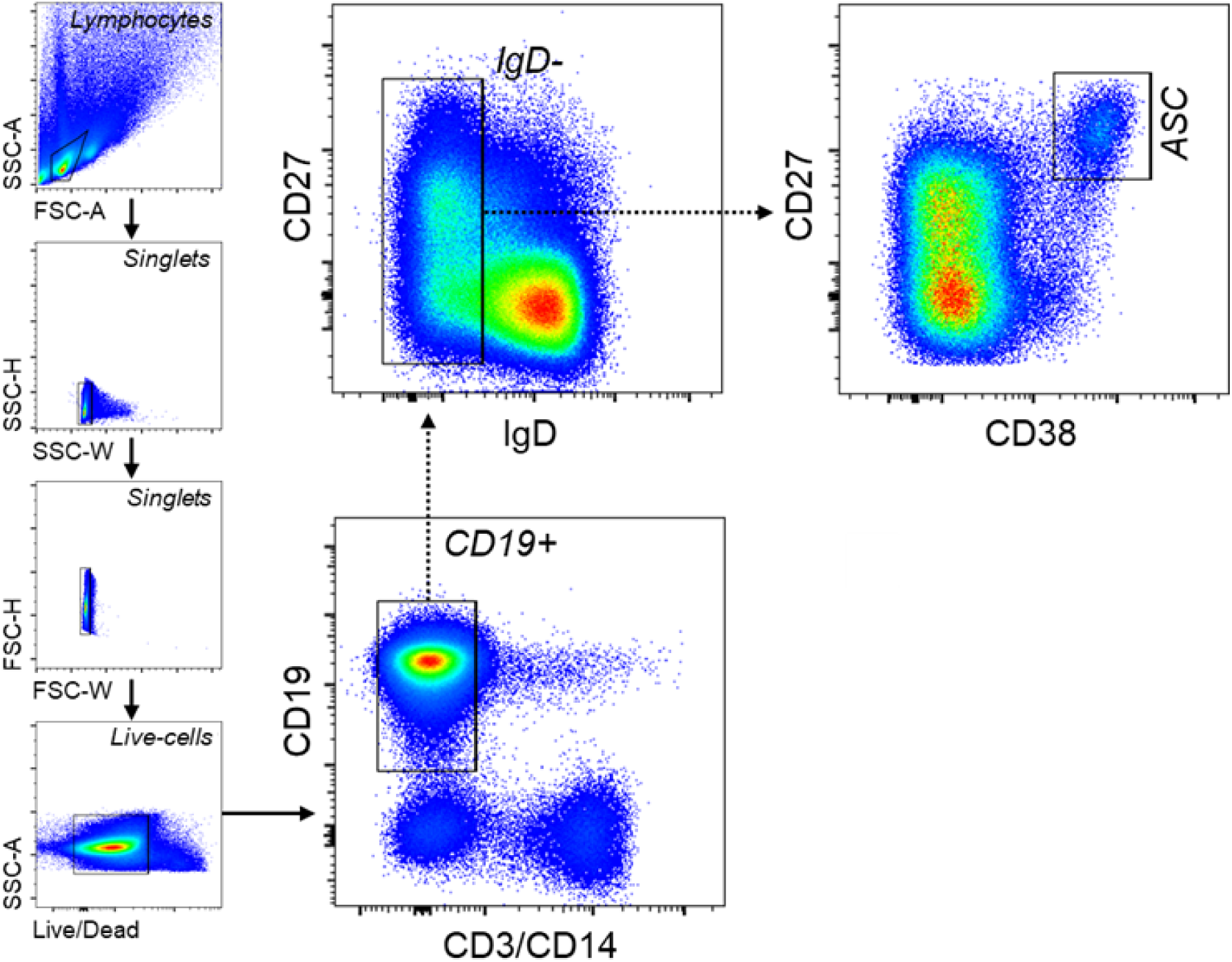
General FACS gating strategy used for sorting blood ASC. PBMC were first gated for lymphocytes, singlets, and viable cells (based on their FSC/SSC and Live/Death properties). CD3 and CD14 were then used as dump markers to capture CD19^+^ and CD19^−^B cell populations. Subsequent sub-gating using CD38 versus CD27 on the IgD^−^ fraction (of CD19^+^ population) allows for sorting for blood ASC (CD27^hi^CD38^hi^). See **Methods** for antibody panels.

**Suppl. Fig. S2.**
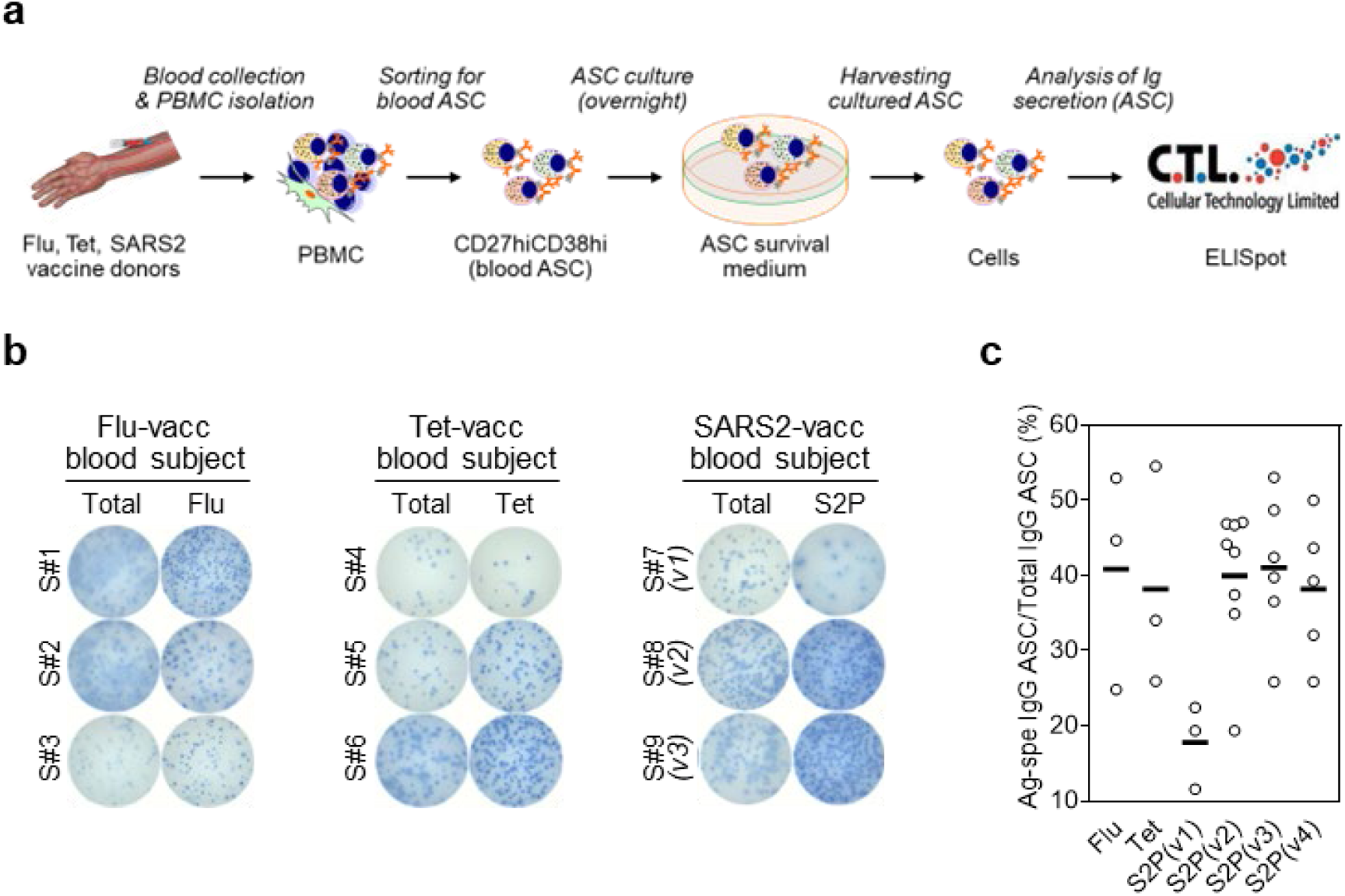
Assessment of Ag-specific ASC and validation of Ag specificities with blood ASC. (**a**) Summary of the techniques and the experimental design. From the cultures of blood ASC, the cells were collected and ELISpot-quantitated for validating Ag specificities. (**b**) Representative ELISpot scanned images shown. Blood ASC from subjects at the peak (ie 5-7 days post-vaccine) assayed for Flu-, Tet-, and S2P-specific IgG. The numbers of input ASC that were incubated were ∼894, ∼1,124, and ∼796 (Total), and ∼4,471, ∼4,496, and ∼2,388 (Flu-specific) for S#1, S#2, and S#3, respectively (far left); ∼1K, ∼1K, and ∼1K (Total), and ∼3K, ∼4K, and ∼4K (Tet-specific) for S#4, S#5, and S#6, respectively (left); ∼712, ∼1,415, and ∼1,386 (Total), and ∼2,139, ∼4,245, and ∼5,544 (S2P-specific) for S#7, S#8, and S#9, respectively (right). (**c**) Each circle represents an individual vaccinee (far right). S, subject; K, 1,000; vacc, vaccinated; Flu, influenza; Tet, tetanus; v, (SARS-CoV-2 mRNA) vaccine dose. All ASC assayed at day 1 in culture.

**Suppl. Fig. S3.**
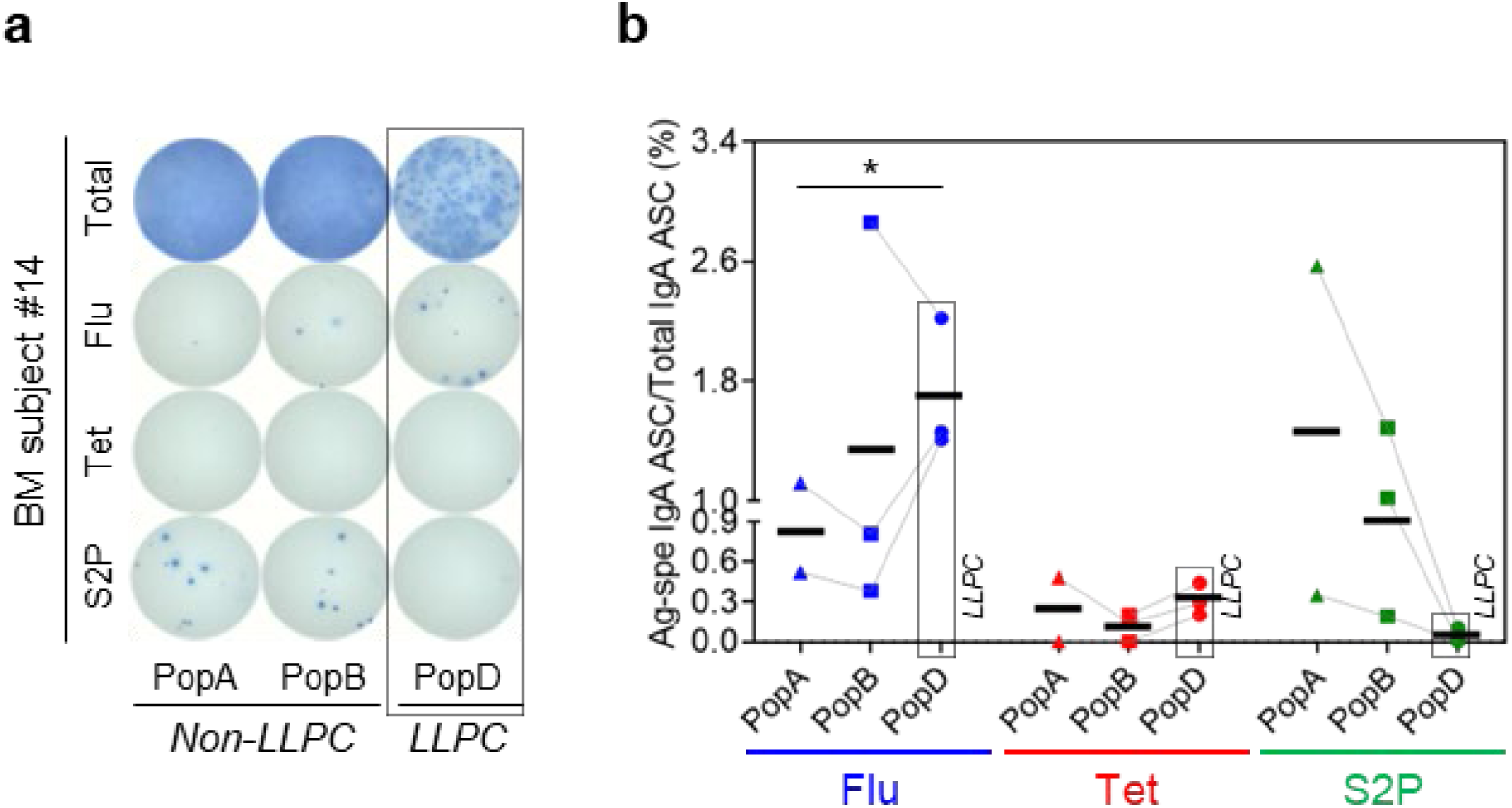
Exclusion of SARS-CoV-2-specific BM IgA LLPC in COVID-19 mRNA vaccinees. (**a**) Representative ELISpot scanned images. The numbers of input ASC that were incubated were ∼9.6K, ∼3.7K, and ∼1.2K (Total) and ∼58K, ∼22K, and ∼7.2K (Ag-specific) for PopA, PopB, and PopD, respectively (left). Unlike Flu-specific, S2P-specific BM IgA ASC emerged in the CD19^+^ (SLPC) compartment (PopA and PopB). (**b**) Each symbol represents an individual vaccinee. K, 1,000; LLPC, long-lived plasma cell (dotted boxes); Flu, influenza; Tet, tetanus. All ASC assayed at day 1 in culture. All comparisons by unpaired t-test between any two subsets for any Ag showed no statistically significant difference (now shown) except for comparison of PopD vs PopA for total IgA; *\*p <0.05*.

**Suppl. Fig. S4.**
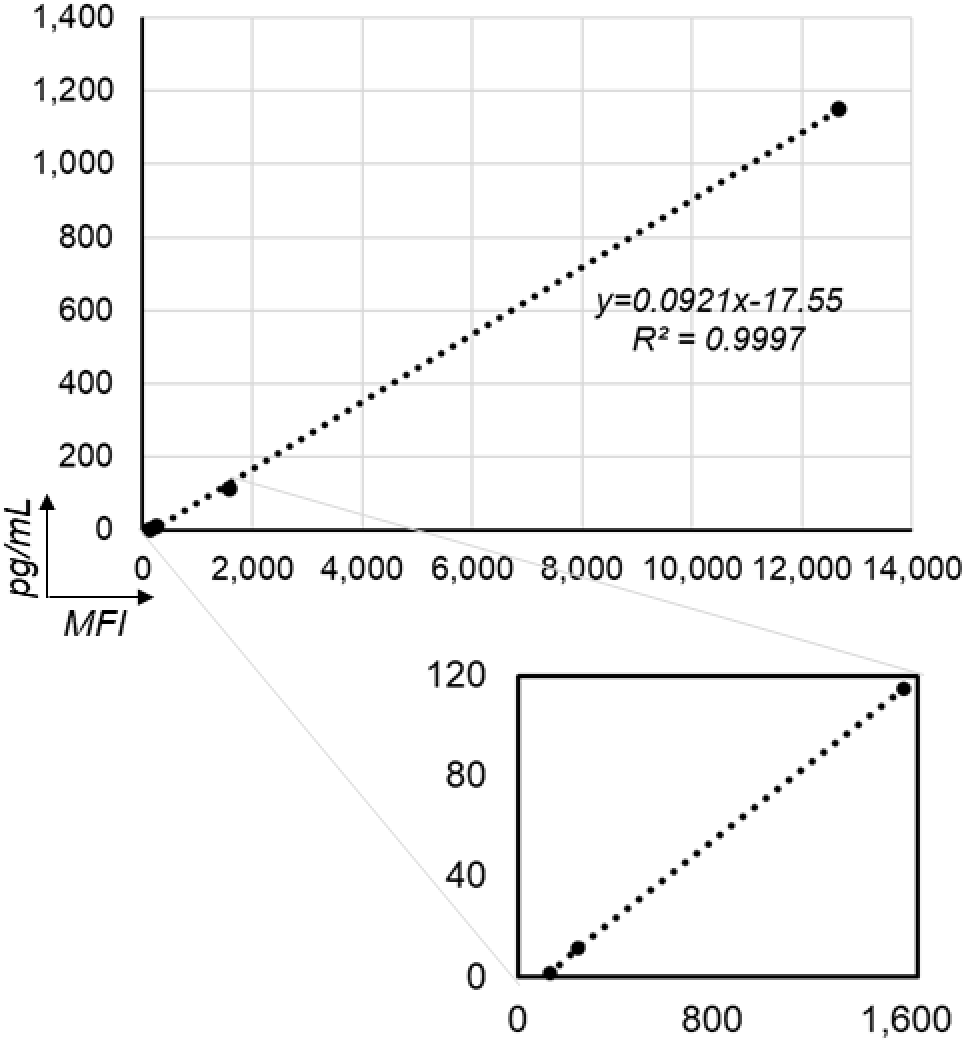
The human IgG standardized concentrations vs MFI values. The displayed equation was used for the purpose of normalization of MFI values generated by MBBA for detection of antibodies in the culture supernatants of BM ASC.

**Suppl. Fig. S5.**
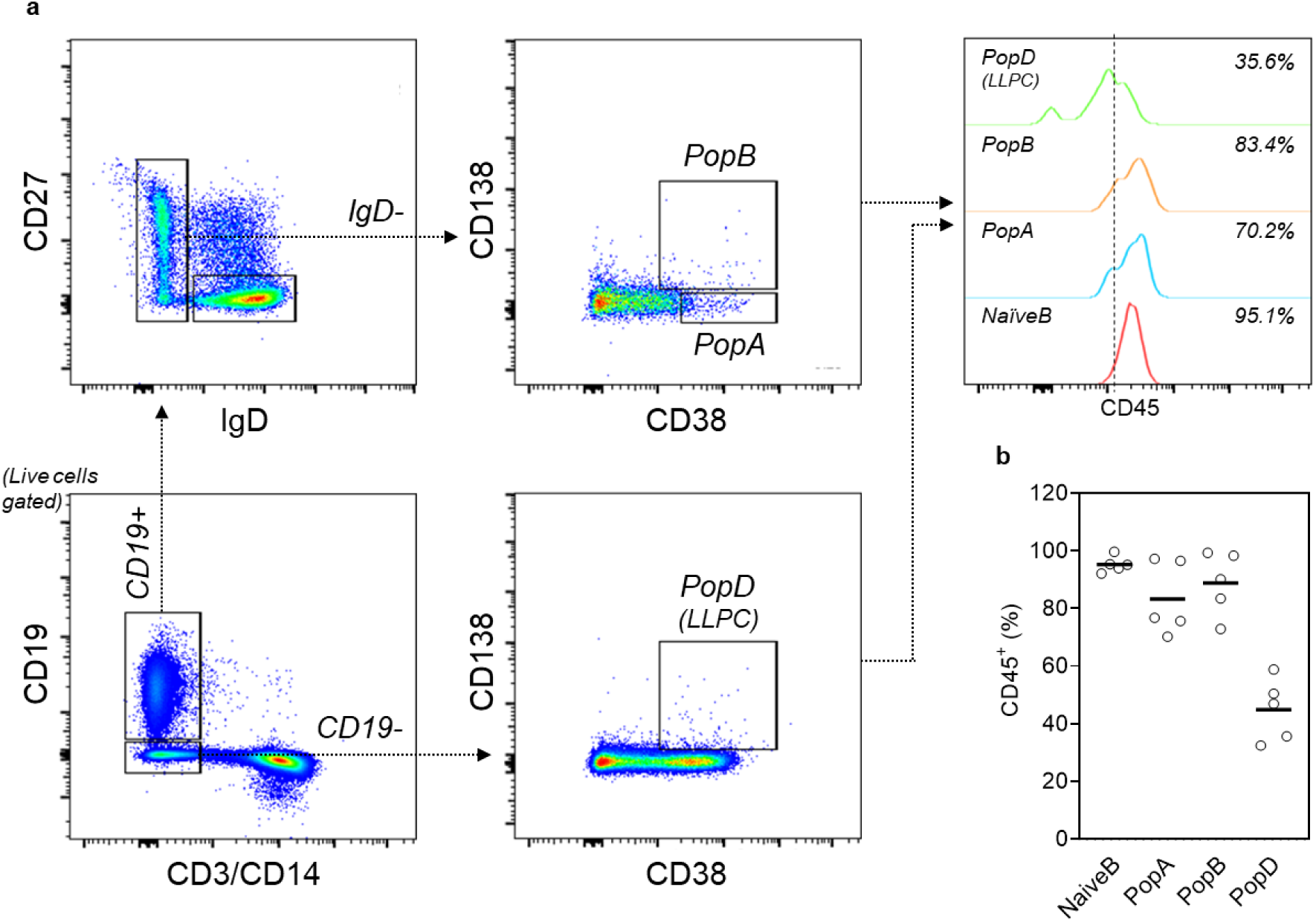
Downregulation of CD45 in LLPC (PopD). (**a**) Representative FACS gating strategy and CD45 staining for BM ASC subsets. For details on BM ASC gating, see Fig. 1b. For the antibody panel, see **Methods**. (**b**) CD45 staining is downregulated in LLPC (PopD). Each circle represents an individual healthy BM donor.

